# The Impact of COVID-19 Lockdowns on the Behavior of Italian Citizens and Particulate Matter 10 and 2.5 Emissions in Lombardy

**DOI:** 10.1101/2020.12.15.20248285

**Authors:** Alessandro Rovetta

**Affiliations:** Mensana srls, Research and Disclosure Division, Brescia, Italy; Redeev srl, Technologic and Scientific Research, Napoli, Italy

**Keywords:** Consumption, COVID-19, Italy, Lockdown, PM10, PM2.5, Sustainability

## Abstract

Italy has been one of the first nations in the world to be heavily affected by the first wave of COVID-19. To date, it is among the first countries for both total cases of contagion and deaths. A wide range of containment measures have been adopted from February to December 2020 to mitigate the pandemic, including total lockdowns across the entire country. This research sets out to evaluate not only how these restrictions influenced Italian citizens’ consumption habits (such as online shopping, smart working and distance learning) but also the impact of lockdowns on the concentrations of particulate matter (PM) 10 and 2.5 in the Lombardy region. In particular, this survey is aimed at investigating the environmental sustainability of the new individual behaviors after the restrictions imposed by the Government in order to quantify their effects on particulate concentrations in Lombardy, the region most damaged by both COVID-19 and air pollution. Various tools and online platforms have been used to collect data, such as Google Trends, web portals providing statistical and demographic information (e.g. AdminStat Italia and ISTAT, which is the National Institute of Statistics), surveys conducted by the Department of Civil Protection, other scientific studies, and the most reliable national newspapers. Technical data on particulate matter was collected from the website of the Regional Agency for the Protection of the Environment (ARPA). To highlight any significant change, the average daily concentrations of PM10 and 2.5 during 2020 in all the provinces of Lombardy were compared with those of the previous year. The comparison between the mean values was made through the t-test. Two values were considered as statistically confident when *t* < 1.5. However, since the real significance of these thresholds is not easily determined, some margins of suspicious confidence have been kept. Finally, using Pearson and Spearman correlations, possible causal correlations between changes in citizens’ behavior and specific key events related to COVID-19 have been dealt with. The P-value threshold was indicatively set at 0.05. Microsoft Excel 2020 and Google Sheets were used as data analysis software. In conclusion, this paper showed a substantial ineffectiveness of total and partial lockdowns in reducing PM10 and PM2.5 concentrations in Lombardy. Furthermore, it has been estimated that COVID-19 has significantly changed the consumption habits of Italian citizens, thus leading to both positive and negative results in terms of sustainability. For instance, a sharp rise in the usage of home delivery services is posing a potential additional threat to the environment at present. At the same time, a positive aspect of this change is the spread of digital literacy, as Italians got quickly acquainted with the most modern technologies for distant learning and smart working, thus paving the way for the establishment of energetically and environmentally sustainable policies throughout the country.

## 1. Introduction

The COVID-19 disease has raged around the world, radically changing habits and feelings of global citizens of all ages and statuses, thus causing deep economic and health crises in all the pandemic stricken-countries (World Health Organization, 2020^1^, Figueroa et al., 2020, Francisco et al., 2020). Among these, Italy is one of the most affected: to date (December 5, 2020), it is the sixth nation in the world for number of deaths from COVID-19, with 1.7 million official cases and 59,000 official deceased (Worldometers, 2020)^2^. However, during the first pandemic wave, lots of media, websites, and environmental protection agencies highlighted a large amount of beneficial effects of the lockdown on pollution and air quality (FASDA, 2020)^3^. In this regard, this research focuses mainly on two aspects: i) the concentrations of particulate matter pollutants with a diameter ranging from 2.5 μm to 10 μm (PM10) and less than 2.5 μm (PM2.5) in the Lombardy region, and ii) the increase in the national demand for home food delivery services, food shopping delivery services, general shopping delivery services, smart working, and distant learning. In particular, the purpose of this paper is to quantify the variation in the concentration of PM10 and PM2.5 as well as evaluate the sustainability of the behaviors adopted by citizens in response to restrictions on personal freedom, containment measures, and lockdowns imposed by the government to face the diffusion of SARS-CoV-2. Because of the multifold aim of this paper, it should be borne in mind that only the Lombardy area was considered for analyzing the variations of PM10 and PM2.5 quantities. The reasons for this are that Lombardy has been the most affected region by both COVID-19 and particulate matter and the deep epidemiological link between the latter and SARS-CoV-2 spread (Rovetta and Castaldo, 2020); this makes it suitable for conducting a reliable investigation and generalizing the results through statistical inference. Conversely, as far as the behavioral response in citizens is concerned, the whole Italian nation has been considered since the threat of contagion was perceived similarly in all regions (Rovetta and Castaldo, 2020). To do this, Google Trends - an online tool provided by Google™-was used as it allows users to search for any query and visualize and quantify the trend over time of the web interest in it in any region of the world. This infoveillance tool is being increasingly exploited by the scientific community for investigating users’ behavior and creating predictive models (Jimenez et al., 2020).

## 2. Methods

### 2.1 Data collection

Google Trends^4^ has been used to explore Italian citizens’ web interest in home food delivery services, food shopping services, shopping services, and smart working software and online learning platforms, during and after the first COVID-19 restrictions and containment measures. Google Trends is a web tool which provides normalized values, called “relative search volumes (RSV)”, ranging from 0 (very low relative interest) to 100 (very high relative interest). All the search parameters and queries are shown in Table 1. PM10 and 2.5 daily averages data were collected from all the measuring stations of every province of Lombardy. To do that, the Regional Agency for Environmental Protection (ARPA) official website^5^ was used. Data on COVID-19 cases and deaths was collected from the Department of Civil Protection official website^6^. Meteorological data was collected from the website “Il Meteo.it”^7^.

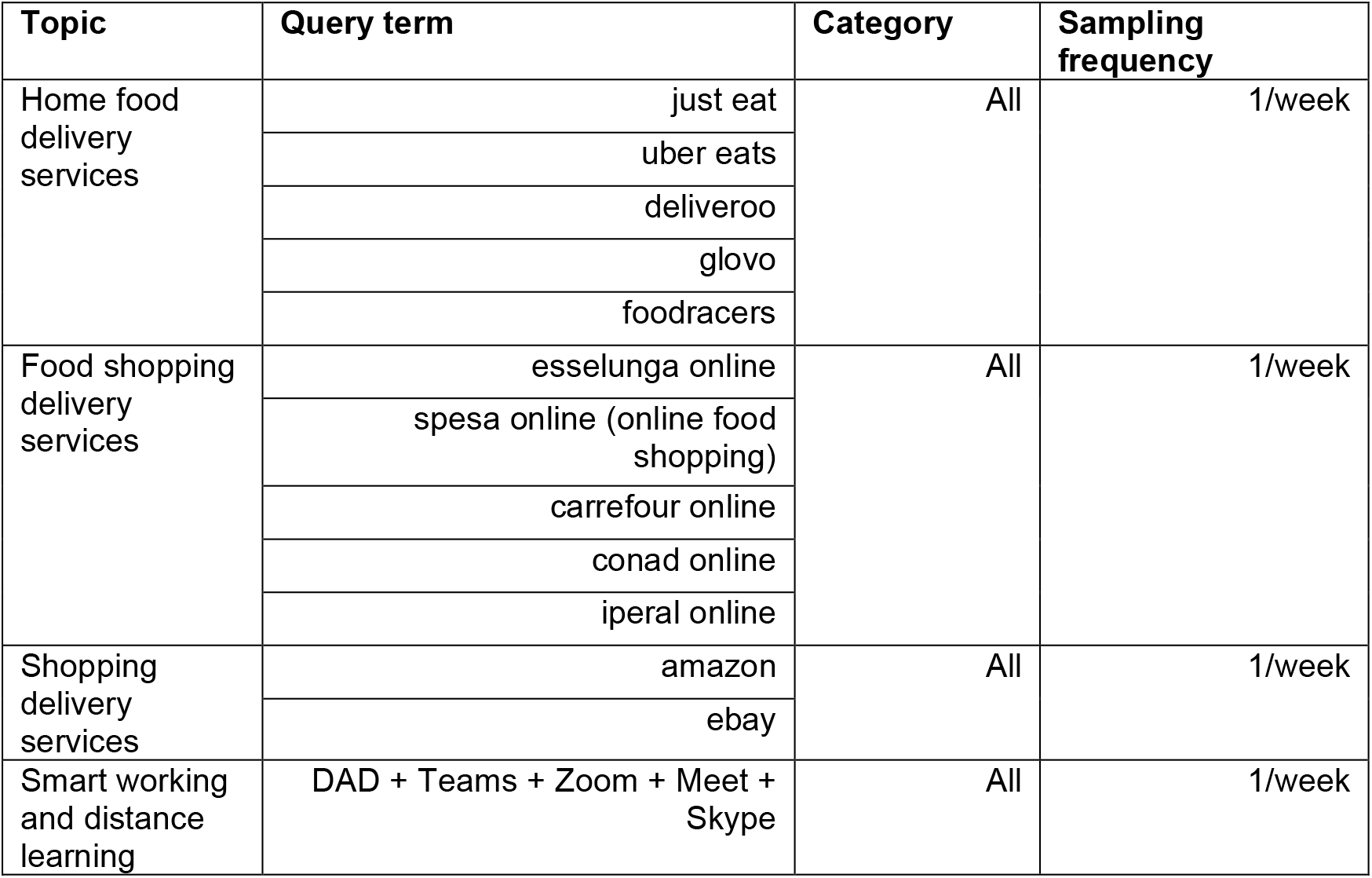

**Table 1.** Weather differences between 2019 and 2020. AT = average temperature, T min = minimum temperature, T max = maximum temperature, HR = relative humidity, Ws = wind speed.

| 2020 vs 2019 |  | Period 1 (1 March - 18 May) |  |  |  |  | Period 2 (1 January - 19 November) |  |  |  |  |
| --- | --- | --- | --- | --- | --- | --- | --- | --- | --- | --- | --- |
|  |  | AT | T min | T max | HR | W | AT | T min | T max | HR | Ws |
| Bergamo | t-test | 2.3 | 2.3 | 1.8 | -1.3 | -2.2 | -0.2 | -0.1 | -0.3 | 0.5 | -1.3 |
| Brescia | t-test | 1.2 | 0.2 | 1.6 | 0 | -1.7 | -0.7 | -0.8 | -0.7 | -0.8 | -2.1 |
| Como | t-test | 1.7 | 1.8 | 1.5 | 1.6 | -2.8 | -0.2 | 0.2 | -0.1 | 1.9 | -2.6 |
| Cremona | t-test | 2.1 | 1.8 | 1.8 | -2.3 | -1.7 | 0.1 | 0.4 | -0.1 | -2.8 | -1.4 |
| Lecco | t-test | 2.3 | 2.3 | 1.8 | -1.3 | -2.2 | -0.2 | -0.1 | -0.3 | 0.5 | -1.3 |
| Mantova | t-test | 2.3 | 1 | 2.1 | -2.7 | -2.1 | -0.3 | -0.4 | 0.1 | 0.4 | -2.1 |
| Milano | t-test | 1.9 | 1.3 | 2 | -1.4 | -2.3 | 0 | 0.2 | 0 | 1 | -2.6 |
| Pavia | t-test | 1.9 | 1.3 | 2 | -1.4 | -2.3 | 0 | 0.2 | 0 | 1 | -2.6 |
| Varese | t-test | 1.7 | 1.8 | 1.5 | 1.6 | -2.8 | -0.2 | 0.2 | -0.1 | 1.9 | -2.6 |

### 2.2 Statistical Analysis

This paper quantifies the impact of COVID-19 restrictions and containment measures on the web queries RSVs and average concentrations of PM10 and 2.5 through the percentage increases *Δ*% = (*x*_1_ − *x*_0_)/*x*_0_. 100 and their related t-test *t* = (*x*_1_ − *x*_0_)/*σ*. All statistical errors *σ* were calculated with the standard propagation formula (Taylor, 1997). Each mean value *x* was reported together with its own standard error of the mean (*SEM*) according to the following modality: *x* ± *SEM* (Taylor, 1997). Data normality was verified both with the z-tests *k*/*σ*_*k*_ < 2 and *s*/*σ*_*s*_ < 2 for kurtosis *k* and skewness *s* and graphically (Tabachnick and Fidell, 1996). For doubtful cases (2 < *z* < 1.5), we also used the Shapiro-Wilk test. When the number of measures *n* was greater than 30, the central limit theorem has been used to validate the t-test when dealing with not normally distributed data (Kwak and Kim, 2017).

#### 2.2.1 Google Trends

We compared the weekly web query trends of 2020 with those of 2019 and 2018, searching for significant discrepancies and focusing on pre-lockdown and the first lockdown period (from 20 February to 18 May 2020). When 2018 and 2019 data series were periodic or stationary, the trends of 2020 queries were compared to those of the previous two years. When 2018 and 2019 data series were aperiodic and non-stationary, anomalous peaks of RSV were highlighted^8^. Two mean values were considered confident only when *t* < 1.5 and not confident only when *t* ≥ 1.9, whereas in the intermediate 1.5 ≤ *t* < 1.9 range non-confidence was interpreted as suspicious. Pearson and Spearman correlations were searched between RSVs and COVID-19 total cases and deaths. The possible causal nature of these relationships was tested by researching the same correlations between the 2019 RSVs and the COVID-19 total cases and deaths during 2020. In order to highlight any hidden correlation, Pearson and Spearman coefficients were used together (Rovetta, 2020). The chosen P-value significance threshold was *α* = .05; however, all correlations with .05 < *P* < .1 were classified as suspicious (Amrhein et al., 2017).

#### 2.2.2 PM10 and 2.5

We compared the average concentrations of PM10 and PM2.5 during 2020 with those of 2019, searching for significant discrepancies and focusing on the first lockdown (period 1: from March 9 to May 18, 2020) and during 2020 (period 2: January 1 to November 19, 2020). Two mean values were considered confident only when *t* < 1.5 and not confident only when *t* ≥ 1.9; in the intermediate 1.5 ≤ *t* < 1.9 range, non-confidence was interpreted as suspicious. Meteorological data - such as minimum, maximum, and average temperatures, % relative humidity, and maximum wind speed-were analyzed according to the same criteria and considering the same timespans. Furthermore, the number of inhabitants and surface area of each province were also reported in the results section, so as to exemplify the coverage of territory of the measuring stations and the demographic importance of all provinces. These data were taken from the official websites of National Institute of Statistics (ISTAT)^9^ and AdminStat^10^.

## 3. Results

Substantial weather changes were identified in Lombardy between 2019 and 2020. In particular, significant or suspected decreases in the maximum wind speed occurred in 100% of cases during period 1 (*Δ*_*av*_% = −18.0 ± 3.3) and in 66.7% of cases during period 2 (*Δ*_*av*_% = −9.2 ± 1.1). In table 1, the t-test results between the meteorological quantities of 2019 and 2020 in 9 of the 12 provinces of Lombardy are shown.

Therefore, the results on the concentrations of particulate matter must be interpreted in the light of the above.

### 3.1 Effect of COVID-19 restrictions on particulate matter concentrations

#### 3.1.0 Lombardy region

During period 1 (i.e. the comparison between the periods 9 March - 18 May 2020 and 2019), significant or suspected increases in the mean concentrations of PM10 and 2.5 were observed in 57% of cases (53/93); significant or suspected reductions occurred only in 3.2% of cases (3/93). In the remaining 39.8% of cases, no significant or suspected changes were observed (37/93). During period 2 (i.e. the comparison between the period 1 January - 19 November, 2020 and 2019), significant or suspected increases in the mean concentrations of PM10 and 2.5 were observed in 20.4% of cases (19/93); significant or suspected reductions occurred only in 6.4% of cases (6/93). In the remaining 73.2% of cases, no significant or suspected changes were found (68/93).

#### 3.1.1 Province of Bergamo

The province of Bergamo has about 1.108 million inhabitants and extends for 2,755 km^2^. In this area, there are 13 measuring stations: 8 for PM10 and 5 for PM2.5. During period 1, 3 significant changes were observed for the average concentrations of PM10 (*Δ*_1_% = +20.0 ± 11.0, *t*_1_ = 2.1; *Δ*_2_% = +28.8 ± 11.7, *t*_2_ = 2.7; *Δ*_3_% = 19.0 ± 10.4, *t*_3_ = 2.1) and 2 worth-noticing variations in the average concentrations of PM2.5 (*Δ*_4_% = +36.5 ± 12.7, *t*_4_ = 3.5; *Δ*_5_ = +23.6 ± 10.8, *t*_5_ = 2.5). 2 suspected cases for PM10 and 1 for PM2.5 were also observed (*Δ*_6_% = +16.4 ± 9.7, *t*_6_ = 1.8; *Δ*_7_% = +14.8 ± 8.6, *t*_7_ = 1.7; *Δ*_8_% = +15.2 ± 11.1, *t*_8_ = 1.5). Therefore, the first lockdown did not cause any significant changes in PM concentrations in 38.5% of cases and caused significant or suspected increases in 61.5% of cases. Considering period 2, all COVID-19 restrictions did not cause significant changes in PM concentrations in 92.3% of cases; only 1 significant change was observed for PM10 (*Δ*_9_% = −13.3 ± 5.3, *t*_9_ = 2.4).

#### 3.1.2 Province of Brescia

The province of Brescia has about 1.264 million inhabitants and extends for 4,786 km^2^. In this area, there are 9 measuring stations: 6 for PM10 and 3 for PM2.5. Only 1 significant change in PM10 average concentration was observed during period 1 (*Δ*% = +23.1 ± 10.5, *t* = 2.4). During period 2, 1 significant change in average PM2.5 concentration (*Δ*_1_% = −10.5 ± 5.5, *t*_1_ = 2.3) and 1 suspected change in average PM10 concentration (*Δ*_2_% = −7.5 ± 5.7, *t*_2_ = 1.5) were observed. Therefore, the first lockdown did not cause significant changes in PM concentrations in 88.9% of cases and caused significant increases in 11.1% of cases. Considering period 2, all COVID-19 restrictions did not cause significant changes in PM concentrations in 77.8% of cases and caused significant or suspected decreases in 22.2% of cases.

#### 3.1.3 Province of Como

The province of Como has about 600,000 inhabitants and extends for 1,279 km^2^. In this area, there are 4 measuring stations: 3 for PM10 and 1 for PM2.5. During period 1, 2 significant changes in average PM10 concentrations were observed (*Δ*_1_% = +25.0 ± 7.8, *t*_1_ = 3.0; *Δ*_2_% = +22.4 ± 7.0, *t*_2_ = 2.7); one suspected change in average PM2.5 concentrations was also found (*Δ*_3_% = +11.8 ± 6.6, *t*_3_ = 1.7). No significant nor suspected changes in PM10 or 2.5 concentrations were observed during period 2. Therefore, the first lockdown did not cause significant changes in PM concentrations in 25.0% of cases and caused significant or suspected increases in PM concentrations in 75.0% of cases. Considering period 2, all COVID-19 restrictions did not cause significant changes in PM concentrations.

#### 3.1.4 Province of Cremona

The province of Cremona has about 360,000 inhabitants and extends for 1,770 km^2^. In this area, there are 10 measuring stations: 6 for PM10 and 4 for PM2.5. During period 1, 3 significant changes were observed for average concentrations of PM10 (*Δ*_1_% = +33.7 ± 12.2, *t*_1_ = 3.3; *Δ*_2_% = +17.1 ± 7.6, *t*_2_ = 2.0; *Δ*_3_% = 16.2 ± 7.8, *t*_3_ = 1.9) and 1 for average concentrations of PM2.5 (*Δ*_4_ = +28.6 ± 7.3, *t*_4_ = 3.4). 2 suspected cases for PM10 and 1 for PM2.5 were also observed (*Δ*_5_% = +12.7 ± 8.0, *t*_5_ = 1.8; *Δ*_6_% = +14.2 ± 8.3, *t*_6_ = 1.6; *Δ*_7_% = +12.6 ± 7.7, *t*_7_ = 1.5). During period 2, only 1 significant change in average PM2.5 concentrations has been observed (*Δ*_1_% = +12.9 ± 5.6, *t*_1_ = 2.2). Therefore, the first lockdown did not cause significant changes in PM concentrations in 30.0% of cases and caused significant or suspected increases in PM concentrations in 70.0% of cases. Considering period 2, all COVID-19 restrictions did not cause significant changes in PM concentrations in 90.0% of cases and caused significant increases in 10.0% of cases.

#### 3.1.5 Province of Lecco

The province of Lecco has about 340,000 inhabitants and extends for 806 km^2^. Here there are 8 measuring stations: 5 for PM10 and 3 for PM2.5. During period 1, 3 significant changes were observed for average concentrations of PM10 (*Δ*_1_% = +46.3 ± 6.5, *t*_1_ = 3.9; *Δ*_2_% = +27.9 ± 7.6, *t*_2_ = 2.6; *Δ*_3_% = 34.6 ± 6.3, *t*_3_ = 3.4) and 2 for average concentrations of PM2.5 (*Δ*_4_% = +30.4 ± 7.8, *t*_4_ = 2.6; *Δ*_5_% = +29.1 ± 8.0, *t*_5_ = 2.5). During period 2, 2 significant changes in average PM10 concentrations (*Δ*_1_% = +11.0 ± 4.9, *t*_1_ = 1.9; *Δ*_2_% = −18.2 ± 7.1, *t*_2_ = 3.5) and 1 for PM2.5 (*Δ*_2_% = −13.1 ± 6.6, *t*_2_ = 2.5) were observed. 1 suspected change was also found (*Δ*_3_% = +9.5 ± 5.2, *t*_3_ = 1.6). Therefore, the first lockdown caused no significant changes in PM concentrations in 37.5% of cases and significant or suspected increases in PM concentrations in 62.5% of cases. As regards period 2, all COVID-19 restrictions caused no significant changes in PM concentrations in 50.0% of cases, significant decreases in 25.0% of cases, and significant or suspected increases in the remaining 25.0%.

#### 3.1.6 Province of Lodi

The province of Lodi has about 230,000 inhabitants and extends for 783 km^2^. In this area, there are 9 measuring stations: 7 for PM10 and 2 for PM2.5. During period 1, significant changes were observed in all measuring stations of PM10 (*Δ*_1_% = +21.9 ± 8.2, *t*_1_ = 1.9; *Δ*_2_% = +29.4 ± 8.6, *t*_2_ = 2.3; *Δ*_3_% = 25.9 ± 7.4, *t*_3_ = 2.5; *Δ*_4_% = +26.0 ± 8.2, *t*_4_ = 2.2; *Δ*_5_% = +23.6 ± 8.5, *t*_5_ = 2.0; *Δ*_6_% = 27.5 ± 7.7, *t*_6_ = 2.4; *Δ*_7_ = +25.6 ± 7.5, *t*_7_ = 2.4) and PM2.5 (*Δ*_8_% = +27.1 ± 9.8, *t*_8_ = 1.9; *Δ*_9_% = +19.4 ± 7.8, *t*_9_ = 1.9). During period 2, 1 significant and 1 suspected changes in average concentrations of PM10 (*Δ*_1_% = +12.4 ± 4.8, *t*_1_ = 2.2; *Δ*_2_% = +8.3 ± 4.6, *t*_2_ = 1.6) and 1 suspected change in average concentrations of PM2.5 (*Δ*_1_% = +10.9 ± 6.2, *t*_1_ = 1.5) were observed. Therefore, the first lockdown caused significant increases in PM concentrations in 100.0% of cases. As regards period 2, all COVID-19 restrictions did not cause significant changes in PM concentrations in 66.7% of cases and caused significant or suspected increases in the remaining 33.3%.

#### 3.1.7 Province of Mantova

The province of Mantova has about 413,000 inhabitants and extends for 2,341 km^2^. In this area, there are 13 measuring stations: 9 for PM10 and 4 for PM2.5. During period 1, 5 significant and 1 suspected changes were observed for average concentrations of PM10 (*Δ*_1_% = +28.4 ± 7.8, *t*_1_ = 2.5; *Δ*_2_% = +25.2 ± 8.1, *t*_2_ = 2.1; *Δ*_3_% = 24.8 ± 7.9, *t*_3_ = 2.2; *Δ*_4_% = +36.3 ± 8.1, *t*_4_ = 2.8; *Δ*_5_% = +30.0 ± 8.3, *t*_5_ = 2.3; *Δ*_6_% = +22.6 ± 9.1, *t*_6_ = 1.8). During period 2, 1 significant and 2 suspected changes in PM10 concentrations were recorded (*Δ*1% = +13.9 ± 4.6, *t*1 = 2.5; *Δ*_2_% = +8.2 ± 4.6, *t*_2_ = 1.6, *Δ*_3_% = 10.1 ± 4.8, *t*_3_ = 1.8). Therefore, the first lockdown did not cause significant changes in PM concentrations in 53.8% of cases and caused significant or suspected increases in the remaining 46.2%. As regards period 2, all COVID-19 restrictions did not cause significant changes in PM concentrations in 84.6% of cases and caused significant or suspected increases in the remaining 15.4%.

#### 3.1.8 Province of Milano

The province of Milano has about 3.26 million inhabitants and extends for 1,575 km^2^. In this area, there are 13 measuring stations: 10 for PM10 and 3 for PM2.5. During period 1, 5 significant changes were observed for average concentrations of PM10 (*Δ*_1_% = +24.3 ± 11.2, *t*_1_ = 2.4; *Δ*_2_% = +26.4 ± 11.4, *t*_2_ = 2.5; *Δ*_3_% = 27.9 ± 12.4, *t*_3_ = 2.7; *Δ*_4_% = −34.2 ± 11.4, *t*_4_ = 3.7; *Δ*_5_% = −21.8 ± 8.4, *t*_5_ = 2.2) and 1 for average concentrations of PM2.5 (*Δ*_6_% = +24.8 ± 13.5, *t*_6_ = 2.1). During period 2, 4 significant changes were observed for average concentrations of PM10 (*Δ*_1_% = +10.8 ± 6.1, *t*_1_ = 1.9; *Δ*_2_% = +11.3 ± 5.9, *t*_2_ = 2.0; *Δ*_3_% = −20.1 ± 3.8, *t*_3_ = 4.9; *Δ*_4_% = +15.3 ± 5.9, *t*_4_ = 2.8) and 1 for average concentrations of PM2.5 (*Δ*_5_% = +18.0 ± 7.6, *t*_5_ = 2.5); 1 suspected change in average PM10 concentrations were also recorded (*Δ*_6_% = +8.3 ± 5.7, *t*_6_ = 1.5). Therefore, the first lockdown did not cause significant changes in PM concentrations in 53.8% of cases, and caused significant increases in 30.8% of cases and significant decreases in 15.4% of cases. As regards period 2, all COVID-19 restrictions did not cause significant changes in PM concentrations in 53.8% of cases, and caused significant or suspected increases in 38.5% of cases and significant decreases in 7.7% of cases.

#### 3.1.9 Province of Monza e Brianza

The province of Monza e Brianza has about 872,000 inhabitants and extends for 406 km^2^. In this area, there are 4 measuring stations: 3 for PM10 and 1 for PM2.5. During period 1, 3 significant changes for average concentrations of PM10 (*Δ*_1_% = +15.7 ± 7.3, *t*_1_ = 1.9; *Δ*_2_% = +28.0 ± 7.2, *t*_2_ = 3.0; *Δ*_3_% = 20.5 ± 6.8, *t*_3_ = 2.8) and 1 for average concentrations of PM2.5 (*Δ*_4_% = +33.0 ± 7.2, *t*_4_ = 3.3) were observed. During period 2, 1 significant change in average concentrations of PM10 (*Δ*_1_% = +12.9 ± 5.4, *t*_1_ = 2.2) and 1 suspected change in average concentration of PM2.5 (*Δ*_2_% = +10.0 ± 5.2, *t*_2_ = 1.8) were recorded. Therefore, the first lockdown caused significant increases in PM concentrations in 100.0% of cases. As regards period 2, all COVID-19 restrictions did not cause significant changes in PM concentrations in 50.0% of cases and caused significant or suspected increases in the remaining 50.0%.

#### 3.1.10 Province of Pavia

The province of Pavia has about 548,000 inhabitants and extends for 2,969 km^2^. In this area, there are 11 measuring stations: 6 for PM10 and 5 for PM2.5. During period 1, 3 significant changes in average concentrations of PM10 (*Δ*_1_% = +24.3 ± 7.4, *t*_1_ = 2.3; *Δ*_2_% = +24.3 ± 7.4, *t*_2_ = 2.3; *Δ*_3_% = 20.4 ± 7.7, *t*_3_ = 2.0) and 3 in average concentrations of PM2.5 (*Δ*_4_% = 32.1 ± 6.4, *t*_4_ = 3.2; *Δ*_5_% = 39.7 ± 7.7, *t*_5_ = 3.0; (*Δ*_6_% = 35.8 ± 6.5, *t*_6_ = 3.4); 2 suspected changes in average concentrations of PM10 and 1 suspected changes in average concentrations of PM2.5 were also found (*Δ*_7_% = +14.7 ± 7.4, *t*_7_ = 1.6; *Δ*_8_% = −13.0 ± 9.4, *t*_8_ = 1.7; *Δ*_9_% = 20.5 ± 10.3, *t*_9_ = 1.5). During period 2, 1 significant change in average concentrations of PM2.5 (*Δ*_1_% = +18.6 ± 5.3, *t*_1_ = 2.6) and 3 suspected change in average concentrations of PM10 (*Δ*_2_% = −11.0 ± 7.4, *t*_2_ = 1.8; *Δ*_3_% = +11.3 ± 6.4, *t*_3_ = 1.5; *Δ*_4_% = +12.2 ± 6.6, *t*_4_ = 1.5). Therefore, the first lockdown caused significant or suspected increases in PM concentrations in 72.7% of cases, suspected decreases in 9.1% of cases, and no significant or suspected changes in 18.2% of cases. As regards period 2, all COVID-19 restrictions did not cause significant changes in PM concentrations in 63.6% of cases, and caused significant or suspected increases in 27.3% of cases and suspected decreases in the remaining 9.1%.

#### 3.1.11 Province of Sondrio

The province of Sondrio has about 182,000 inhabitants and extends for 3,196 km^2^. In this area, there are 6 measuring stations: 4 for PM10 and 2 for PM2.5. During period 1, 3 significant changes for average concentrations of PM10 (*Δ*_1_% = +21.6 ± 6.4, *t*_1_ = 2.5; *Δ*_2_% = +17.7 ± 5.5, *t*_2_ = 2.5; *Δ*_3_% = 27.0 ± 6.4, *t*_3_ = 3.0) and 1 for average concentration of PM2.5 (*Δ*_4_% = +20.5 ± 5.9, *t*_4_ = 2.7); 1 suspected change has occurred for average concentrations of PM10 (*Δ*_5_% = +14.0 ± 6.8, *t*_5_ = 1.7). During period 2, no significant or suspected changes were recorded. Therefore, the first lockdown caused significant increases in PM concentrations in 83.3% of cases and no significant changes in the remaining 16.7%. As regards period 2, no significant or suspected changes were observed.

#### 3.1.12 Province of Varese

The province of Sondrio has about 890,000 inhabitants and extends for 1,198 km^2^. In this area, there are 6 measuring stations: 4 for PM10 and 2 for PM2.5. During period 1, 1 significant change in average concentrations of PM10 (*Δ*_1_% = +21.6 ± 7.6, *t*_1_ = 2.1) and 1 in average concentrations of PM2.5 (*Δ*_2_% = +27.9 ± 7.9, *t*_2_ = 2.4) were observed. During period 2, only 1 suspected change in average PM2.5 concentrations was found (*Δ*_1_% = +11.9 ± 6.4, *t*_1_ = 1.7). Therefore, the first lockdown did not cause significant changes in PM concentrations in 66.7% of cases and caused significant increases in the remaining 33.3%. As regards period 2, all COVID-19 restrictions did not cause significant changes in PM concentrations in 83.3% of cases and caused significant or suspected increases in the remaining 16.7%.

### 3.2 Consumer behavior

#### 3.2.1 Home food delivery services

Italian citizens’ web interest in home food delivery services showed seasonality during 2018 and 2019 (Figure 1). During the first lockdown and pre-lockdown (20 February - 18 May, 2020), the queries “just eat, uber eats, deliveroo, foodracers and glovo” recorded a significant percentage increase if compared with the same period in 2019 (*Δ*_1_% = +30.6 ± 6.5, *t*_1_ = 4.8; *Δ*_2_% = +80.7 ± 58.0, *t*_2_ = 2.9; *Δ*_3_% = +38.7 ± 25.1, *t*_3_ = 5.8; *Δ*_4_% = +269.2 ± 60.5, *t*_4_ = 5.2; *Δ*_5_% = +173.0 ± 44.8, *t*_5_ = 8.3). It must be pointed out that three of these queries (uber eats, deliveroo, and glovo) had already undergone a significant increase between 2018 and 2019. However, the highest RSV peak detected in spring 2020, during the first lockdown, was not present in 2018 and 2019 series (Figure 1). Moreover, such trends have also been heavily influenced in the months following the first wave of COVID-19 (Figure 1). Significant correlations were observed between RSVs and COVID-19 total cases and total deaths (*r*_1_ = .78, *P* < .0001; *r*_2_ = .72, *P* = .0003); nevertheless, such correlations were also identified between the same web queries during 2019 and COVID-19 total cases and deaths.

**Figure 1.**
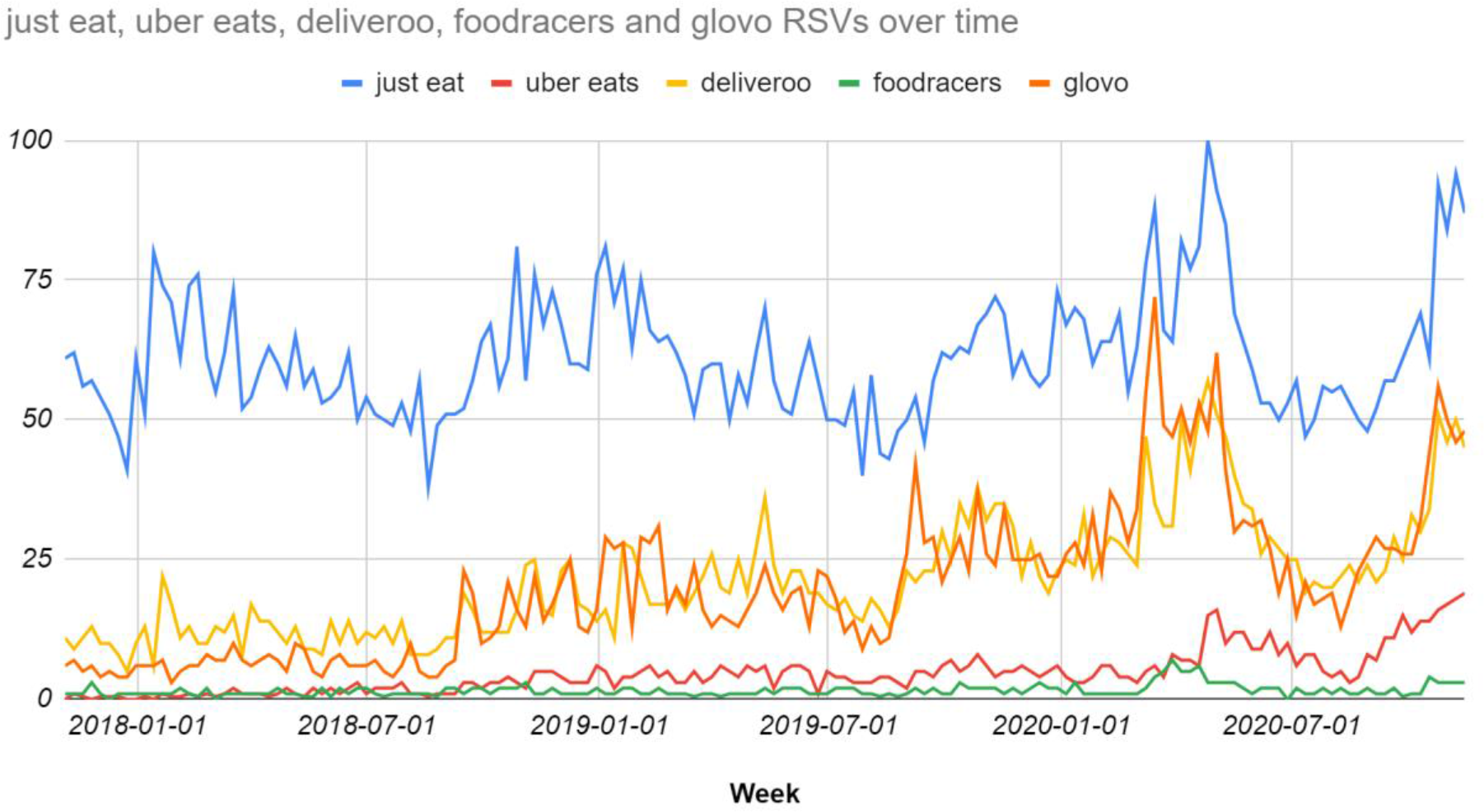
Italian citizens’ web interest in home food delivery services from November 2017 to November 2020.

#### 3.2.2 Online food shopping services and online supermarkets

The web interest of Italian citizens in food shopping services and online supermarkets underwent a significant percentage increase during the first lockdown compared to the same trend during 2018 and 2019 (Figure 2). Indeed, the following queries including names of some of the commonest Italian supermarket chains (i.e. carrefour online, conad online, iperal online, esselunga online) alongside more general ones about online food shopping (i.e. spesa online + supermercato online + supermercati online) recorded the following percentage increases: *Δ*_1_% = +861.9 ± 293.8, *t*_1_ = 3.4; *Δ*_2_% = +2593.8 ± 725.9, *t*_2_ = 3.4; *Δ*_3_% = +1100.0 ± 324.6, *t*_3_ = 2.9; *Δ*_4_% = +1358.3 ± 450.4, *t*_4_ = 2.9; *Δ*_5_% = +184.6 ± 65.4, *t*_5_ = 3.4. Moreover, the interest in these queries remained relatively high during the rest of 2020 compared to those of the previous two years, growing again with the arrival of the second period of restrictions (Figure 2). Significant hidden correlations were observed between RSVs and COVID-19 total cases and deaths (*R*_1_ = .49, *P* < .035; *R*_2_ = .46, *P* = .049); nevertheless, such correlations were also identified between the same web queries during 2019 and COVID-19 total cases and deaths.

**Figure 2.**
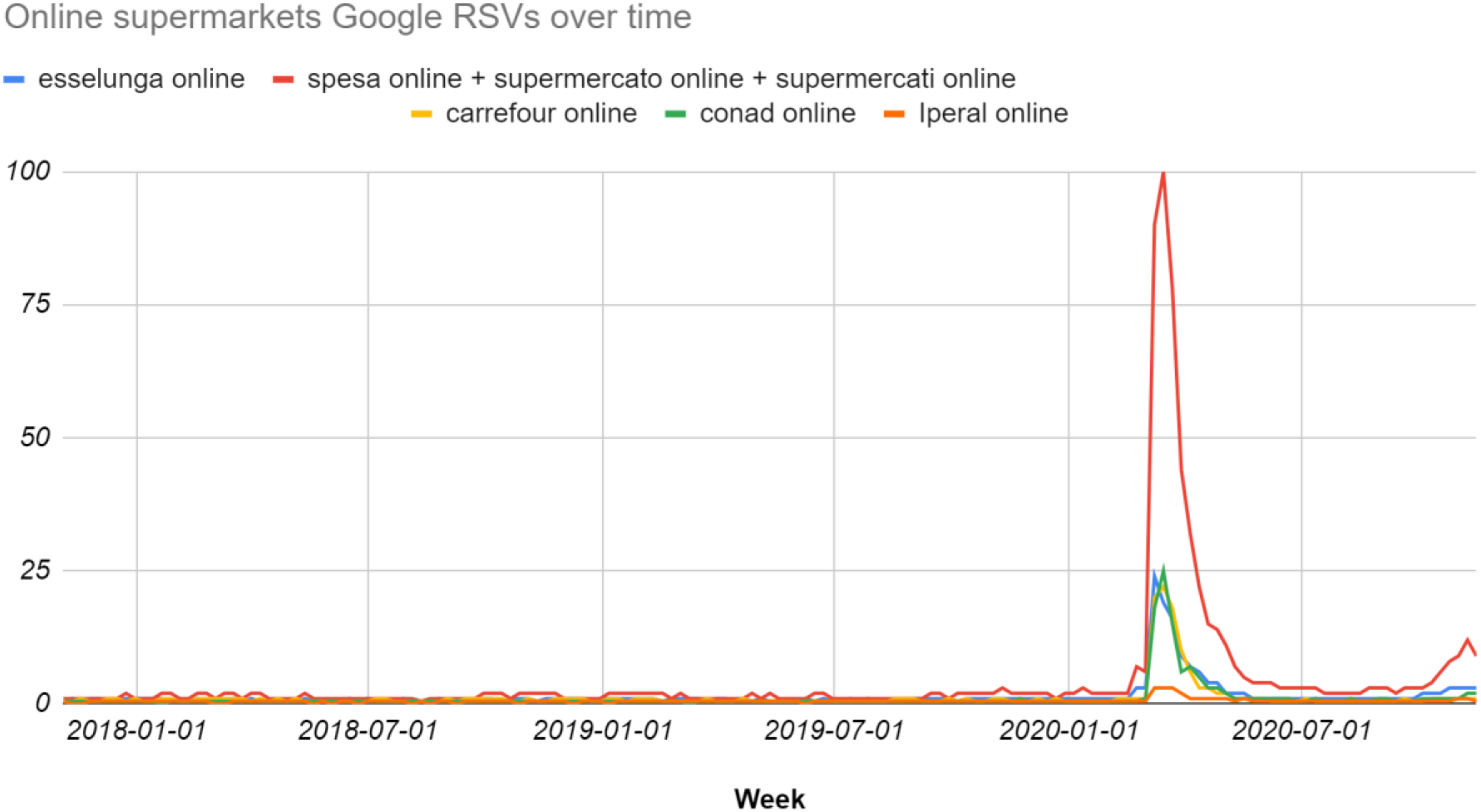
Italian citizens’ web interest in supermarket delivery services from November 2017 to November 2020.

#### 3.2.3 Online shopping

The web interest of Italian citizens in home delivery services showed seasonality during 2018 and 2019 (Figure 3). During the pre-lockdown and first lockdown (20 February - 18 May, 2020), the query “amazon” recorded a significant percentage increase if compared with the same period in 2019 (*Δ*% = +22.1 ± 4.0, *t* = 6.2). This increase continued throughout the spring and early summer to varying degrees (Figure 3). The query “ebay” also underwent an anomalous rise during the first lockdown period, reaching the annual RSV peak despite what happened in 2018 and 2019 (Figure 3). Only low and non significant correlations were identified between such queries and COVID-19 total cases and deaths (*R, r* < 0.3, *P* > .23).

**Figure 3.**
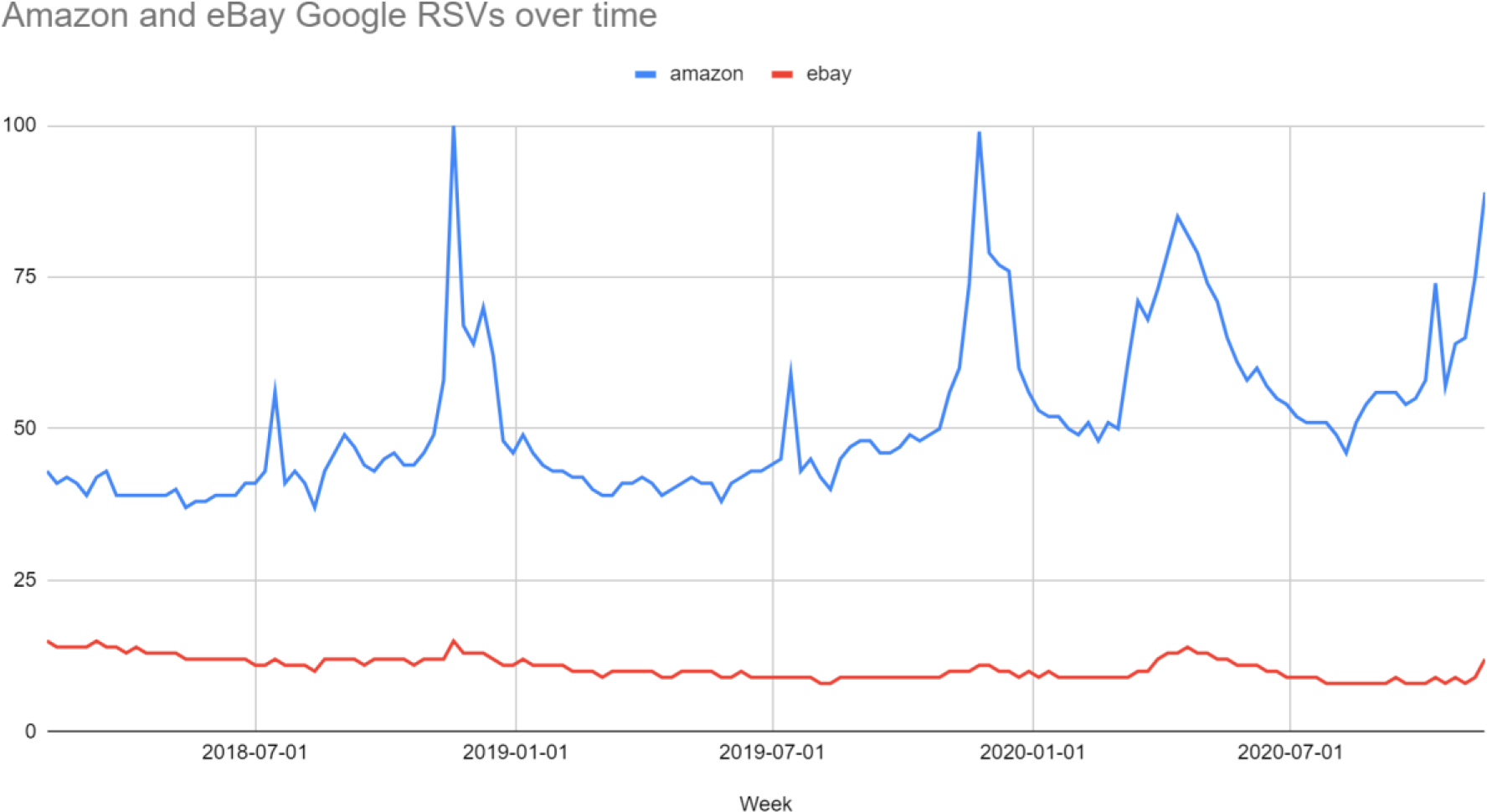
Italian citizens’ web interest in shopping delivery services from November 2017 to November 2020.

#### 3.2.4 Smart working and distance learning

The web interest of Italian citizens in smart working and distance learning platforms such as Google Classroom, Teams, Zoom, Google Meet, and Skype, underwent a drastic surge during and after the first lockdown (*Δ*% = + 1190.8 ± 133.3, *t* = 8.8 during period 1 and *Δ*% = + 632.7 ± 78.8, *t* = 8.2 during period 2). Among all regions, Calabria is the one that has shown the highest interest in these technologies with an *RSV* = 100. The other regions had an *RSV* in the interval [35, 50] i.e. they showed a more similar interest (*SD* 4.3).

## 4. Discussion

Several studies have been conducted on the impact of the lockdown on Lombardy’s air quality (Cameletti, 2020, Piccoli et al., 2020, Putaud et al., 2020, and Collivignarelli et al., 2020). However, not all results are fully concordant: in fact, while Putaud et al. claim that they have not observed any significant change in PM10 concentrations in Milan, Collivignarelli et al. argue that the lockdown had a positive effect on them, as well as on PM2.5 and other air pollutants. In this context, Cameletti highlights how a reduction in particulate matter emissions does not necessarily imply a reduction in its concentrations, underlining the complexity in the analysis of these types of phenomena. Such evidence could help explain the discrepancy found between previous authors and between the various reports from environmental protection agencies and universities. Another point to be taken into account is that these differences could also be explained by the different approaches adopted in the aforementioned studies, since some have used predictive models inherent exclusively to 2020 while others have compared the concentrations of pollution in the current year with those of previous years. This paper falls into the second category, as it investigates the differences in average concentrations of PM10 and PM2.5 between specific periods during 2019 and 2020. Piccoli et al. showed that, as Milan registered similar temperatures during February and March in both 2019 and 2020, such data are suitable for a comparative analysis. This supports the assumption of the absence of unusual weather regimes. However, an evident reduction in wind speed was highlighted in this paper which certainly contributed to the increase in concentrations of particulate matter across the Lombardy region. Moreover, non-uniform and significant variations in temperature and relative humidity may have further biased the estimate of particulate matter emissions in an unpredictable manner. Nonetheless, the most important result of this paper is that even a total lockdown strategy would be absolutely insufficient to reduce the concentrations of PM10 and PM2.5 in a sustainable way. Similar results were found by Donzelli et al. in Tuscany (Donzelli et al., 2020). This again calls into question the role of motor vehicles in the production of this type of air pollution. Abu-Allaban et al. showed that the fractional contributions of motor vehicles to ambient PM in urban areas in the USA were in the ranges 20-76% and 35-92% for PM2.5 and PM10, respectively. The Italian National Research Council (CNR)^11^ measured a reduction in road traffic of 48-60% in Italian cities which was obviously not followed by an equal reduction in particulates, thus confirming the great variance of the above ranges. Therefore, this research suggests that industries, private routinary activities, and ambiental factors, could contribute very significantly to air pollution in Lombardy. Moreover, the demand for online delivery and shopping services has dramatically increased both during and after lockdowns. All of these e-commerce services-including Amazon-can have a very negative environmental impact, especially when providing fast delivery times: indeed, not only the emissions of the transport vehicles but also the plastic packaging widely used by the companies do contribute to environmental pollution^12,13^ (CNN, 2019, SCSC, 2019). In this regard, Amazon claims to be planning a sustainable delivery strategy^14^ (Amazon, 2020): considering the current condition, it is high time the company was faithful to its words. Furthermore, it would be advisable that states and organizations all over the world impose more stringent and effective criteria for monitoring the individuals’ environmental impact, as far as their administrative policies are concerned. Another aspect to be considered is that Google Trends has proved to be an effective infoveillance tool for this type of scientific survey, since the growing web interest in food delivery services has been met by a real increase in the orders actually placed by Italian users (AGI, 2020)^15^. Finally, smart working and distance learning could be very valid alternatives in terms of environmental impact as they do not involve travelling or excessive use of materials such as textbooks, notepads, notebooks, chalks, markers, etc. Conversely, this leads to an increase in household consumption of electricity and heat, especially during the winter. Nonetheless, it should be taken into account that such consumptions were inevitable even when people work in their own work structures and private houses in Italy do not often have the same energy class systems (ABC Costruzioni, 2018). Moreover, there are many challenges to be faced before taking this path (The Planet Mark, 2016, Hodder, 2020, Sarti et al., 2020) and the effectiveness of online lessons is still a hotly-debated topic among parents, teachers and psychologists (Daniel, 2020, Mukhtar et al., 2020, Abbasi et al., 2020, Abuhammad, 2020). What is certain is that the population has had to forcibly learn to use some of the most modern technologies and techniques of file sharing, databases, and online communication software and tools; thus, in the future, it will be easier and less impacting at an emotional-psychological level to insert smart working and online learning methods in everyday life, once the right compromise between the psychosocial health safeguard and environmental protection will be reached. Finally, the correlations found between COVID-19 cases and deaths, and web searches related to home delivery services in 2019 and 2020, could prove how the virus has spread easier in regions with more frequent trade relations.

### 4.1 Strengths and limitations

As far as the author knows, this is the first study that compares PM10 and PM2.5 concentrations during 2019 and 2020 in Lombardy including data from all the measuring stations of all its provinces also quantifying their meteorological differences. Furthermore, this study demonstrates a substantial reliability of Google Trends in estimating the demand for e-commerce and home delivery services of Italian web users as well as the ineffectiveness of lockdowns in the reduction of PM10 and PM2.5 concentrations in the most populated Italian region. However, some limitations must be considered: the search for statistical correlations cannot provide details on any possible causal nature of the phenomena investigated. The web interest of Google users is not certain to reflect the general interest of all Italian citizens. In some provinces, the number of measuring stations could be insufficient to represent in detail the overall trend of PM10 and PM2.5 concentrations. Finally, some weather differences may have biased the comparison between the particulate emissions during 2019 and 2020.

## 5. Conclusions

This study shows that even a forced total lockdown strategy, capable of reducing road traffic in Italian cities by 48-60%, was absolutely insufficient to reduce the concentrations of PM10 and PM2.5 in the Lombardy region. Environmental factors, industries, and individual routinary activities, can therefore have non-marginal or even preponderant roles in the production of this type of pollution. At the same time, COVID-19 has shown that citizens and the government are not yet ready to manage policies that support smart working or distance learning. Furthermore, the increase in demand for home delivery services and online shopping during and after the lockdowns must draw further attention from government authorities on the environmental issue. In conclusion, we can state that COVID-19 has drastically changed various habits of Italian citizens. Among the positive aspects of this change there was the learning of new technological methods and tools for online collaboration and remote communication via the Internet. This can be a starting point for the integration of policies aimed at preserving both the mental and physical health of citizens and the environment on which they depend.

## Data Availability

All data are available in the maunscript

## Footnotes

1 https://www.who.int/news/item/13-10-2020-impact-of-covid-19-on-people’s-livelihoods-their-health-and-our-food-systems

2 https://www.worldometers.info/coronavirus/

3 https://www.fasda.it/effetti-quarantena-ambiente/

4 https://trends.google.com/trends/

5 https://www.arpalombardia.it/Pages/Aria/Richiesta-Dati.aspx

6 https://github.com/pcm-dpc/COVID-19

7 https://www.ilmeteo.it/portale/archivio-meteo/Lombardia

8 Anomalous peaks are defined as upward trends possibly related to lockdowns or other key events as these were not present in the previous two years trends.

9 https://www.istat.it/

10 https://ugeo.urbistat.com/AdminStat/it

11 https://www.cnr.it/it/comunicato-stampa/9702/l-inquinamento-in-italia-durante-il-lockdown

12 https://edition.cnn.com/2019/07/15/business/fast-shipping-environmental-impact/index.html

13 https://sustainability.aboutamazon.com/environment/packaging-and-products

14 https://sustainability.aboutamazon.com/environment/packaging-and-products

15 https://www.agi.it/economia/news/2020-04-22/fase-2-coronavirus-amazon-spedizioni-8411103/

## Notes

### Competing Interest Statement

The authors have declared no competing interest.

### Funding Statement

None received

